# Triangulating evidence for the causal impact of single-dose zinc supplement on glycemic control for type-2 diabetes

**DOI:** 10.1101/2021.12.17.21267964

**Authors:** Zhiyang Wang, Carine Ronsmans, Benjamin Woolf

## Abstract

**Background:** Although previous studies suggested the protective effect of zinc for type-2 diabetes, the unitary causal effect remains inconclusive.

**Objective:** We investigated the causal effect of zinc as a single intervention on glycemic control in type-2 diabetes patients, using a systematic review of RCTs and two-sample Mendelian randomization (MR).

**Methods:** Four outcomes were identified: fasting blood glucose/fasting glucose, hemoglobin A1c (HbA1c), homeostasis model assessment of insulin resistance (HOMA-IR), and serum insulin/fasting insulin level. In the systematic review, four databases were searched up to June 2021. Results were synthesized through the random-effects meta-analysis. Single nucleotide polymorphisms (SNPs) that are independent and are strongly related to zinc supplements were selected from MR-base to perform the two-sample MR with inverse-variance weighted (IVW) coefficient.

**Results:** In the systematic review, 14 trials were included. The zinc supplement led to a significant reduction in the post-trial mean of fasting blood glucose (mean difference (MD): - 26.52, 95%CI: -35.13, -17.91), HbA1C (MD: -0.52, 95%CI: -0.90, -0.13), and HOMA-IR (MD: -1.65, 95%CI: -2.62, -0.68), compared to the control group. In the two-sample MR, zinc supplement with 2 SNPs associated with lower fasting glucose (IVW coefficient: -2.04, 95%CI: -3.26, -0.83), but not specified type-2 diabetes.

**Conclusion:** Although the study was limited by the few trials (review) and SNPs (two-sample MR), we demonstrated that the single zinc supplementary improved glycemic control among type-2 diabetes patients with causal evidence to a certain extent.

## 1. Introduction

Type 2 diabetes (T2D) is a chronic disease in that patient’s body dose not normally respond to insulin, called insulin resistance. Impaired insulin sensitivity reduces the absorption and reserve of insulin in organs, decreasing serum insulin and increasing blood sugar levels^1^. Its common symptoms include polydipsia, fatigue, and unintended weight loss. Individually, diabetes causes lower self-evaluation of physical and mental health, inducing psychological comorbiditiy^2^. Globally, the age-standardized incidence rate increased by 3.23% to 279.13 per one million from 2007 to 2017 and the disability-adjusted life years increased by 5.07%^3^. The growing number of T2D cases leads to a heavier burden for global health.

T2D brings multiple challenges to medicine, primary care, and the economy. Four-fifths of diabetes patients currently live in low- and middle-income countries (LMIC), and the susceptibility to infection and worse outcomes such as hepatitis B and HIV would also increase^4^. In India, nearly half of patients did not receive a proper diagnosis of T2D, and the hidden progression of complications was more threatening^5^. Moreover, patients faced substantial cost-of-illness and lower employment chances, and the government’s health expenditure on T2D was projected to incline^6, 7^. It is therefore critical to find a cost-effective and accessible intervention to address the threat of T2D. Zinc is a common commercial supplement, which makes it a good candidate solution.

Zinc supplementation was demonstrated to have a protective effect on insulin and metabolism in the hyperglycemic environment in animal models^8–10^. The zinc ion plays an essential role with insulin in the pancreatic β cell, activating multiple cell signaling cascades^11^. Zinc ions coordinate six insulin monomers (hexamerization), which enhance the stability of insulin and the storage capacity of the insulin-secreting vesicles^12–14^. Moreover, zinc is known for its antioxidative property as a cofactor of the important antioxidative enzyme, which may reduce the lipid peroxidation and development of insulin resistance in diabetes mellitus^15–17^.

There are many randomized controlled trials (RCT) which evaluate the impact of zinc or co-supplements on various types and progression of diabetes^18, 19^. One meta-analysis published in 2019 included 36 studies and found that zinc supplements in both single factor and co-supplements significantly reduced glycemic indicators such as fasting glucose^20^. In a 2021 meta-analysis of 27 studies, researchers suggested that low-dose and long-duration single zinc supplements had a beneficial impact on many T2D and cardiovascular disease risk factors^21^. The multi-nutrients intervention in the previous review incorporated the interaction between nutrients, so that we may not get a valid estimate of zinc’s own effect. For example, excess amounts of iron and calcium compromised the zinc level and inhibited its bioaccessibility, and the interaction mechanism within more micronutrients remained complicated^22, 23^. Both studies did not limit the type of diabetes or even included healthy participants at high risk so that the real effect of the single zinc supplement on prevalent T2D patients was masked and needed more emphasis.

Mendelian randomization (MR) also helps us to evaluate the causality between single zinc supplements and diabetes. MR is regarded as a “natural experiment”, which leverages the random inheritance of genetic variants to approximate the random allocation of the participants to a modifiable exposure which the variants are robustly associated with^24–27^. Because genes are inherited at birth, MR estimates are interpreted as the lifetime effect of the exposure, while RCT studies can only provide the effects over shorter periods of time. This innovative method has been used in previous nutrition and metabolism studies to obtain an unbiased assessment of nutritional status to the outcome of interest. Researchers notably demonstrated the association of long-term testosterone exposure with health outcomes by MR^28^. Collaborating with RCT, MR could help to strengthen the causal estimation, particularly in cumulating exposure inside the body or capturing a sensitive period of the life course^29^

In this study, we aimed to systematically assess the possible causal inference about the association between zinc supplements and glycemic control among T2D patients, using a systematic review of RCTs and two-sample MR.

## 2. Method

### 2.1 Systematic review

#### 2.1.1 Protocol and ethnics

This meta-analysis followed the Preferred Reporting Items for Systematic Reviews and Meta-analyses (PRISMA) reporting guideline^30^. This study received ethics approval from the London School of Hygiene and Tropical Medicine MSc Research Ethics Committee. The methodology of the review and analysis was approved in advance by the LSHTM epidemiology MSc course directors.

#### 2.1.2 Eligibility criteria

The study eligibility criteria were specified using Population, Intervention, Comparison, Outcomes, and Study (PICOS) frame^30^.

##### Inclusion criteria

♦ Study population: Any human participants with type-2 diabetes. There was no restriction on the demographic characteristics.
♦ Intervention: Zinc administration as the single supplement intervention.
♦ Comparison: Use of placebo.
♦ Outcome: Primary outcomes included fasting blood glucose, hemoglobin A1C (HbA1c), homeostatic model assessment for insulin resistance (HOMA-IR), and serum insulin level. Secondary outcomes included any other quantitative outcomes related to diabetes control.
♦ Study design: Any randomized controlled trial evaluating the association between zinc intervention and glycemic control.

##### Exclusion criteria

♦ Full-text not available or accessible.
♦ Co-supplement with other nutrition would not be included to concentrate on the zinc- only effect.

#### 2.1.3 Information sources and search strategy

PubMed, CINAHL Plus, EMBASE, and Web of Science were searched from the establishment time of each database to June 2021. We also checked the references and citations of all included studies for eligibility criteria. The search strategy was developed by and modified from the previous systematic review^20^. The terms were structured according to each databasès guidelines and searched by titles and abstracts. The detail was presented in supplementary material 1.

#### 2.1.4 Selection process

The abstracts and titles were screened by the eligibility criteria. The first unblinded reviewer screened all the citations and the second one checked a random 10% sample of all citations, due to the limitation of human resources. The percentage agreement and the Gwet AC (agreement change adjusted) between the two reviewers were calculated to test the interrater reliability^31^.

#### 2.1.5 Data extraction and item

The standard form for intervention reviews for RCTs developed by the Cochrane Collaboration was used to exact the data^32^. The extraction process was conducted twice to minimize bias or entry error. The specific data items are shown below:

♦ Eligibility criteria: inclusion and exclusion criteria
♦ Study design (parallel or crossover), assignment of each arm, and analysis plan (intention-to-treat or per-protocol)
♦ Participants: the total number of randomized and analyzed participants, numbers in each arm (pre-trial and post-trial), geographic information, and demographic characteristics.
♦ Intervention and control arms: formulation and dosage of Zinc, administration method, description of the control arm (placebo), and study duration.
♦ Results: the measure of effect (means, mean difference, or change score), standard error of effect measure, statistical significance, and any other results such as odds ratio.
♦ Information about risk-of-bias: any information causing bias, such as missing participants.

#### 2.1.6 Effect measure

The primary measure was the post-trial mean difference (MD) with standard error (SE) for each outcome. The change scores are the change of mean from pre-trial to post-trial in each arm so that the difference of change scores between zinc intervention and control was our secondary effect measure. Converting units or the standardized mean difference (SMD) were used to get the uniform scale^33^.

#### 2.1.7 Synthesis methods

The overall meta-analyses and the subgroup meta-analyses were conducted to synthesize the results. The subgroup analyses were stratified by studies that clearly stated the participants were T2D patients and that did not specify the participant’s diabetes type.

The Cochran-Q χ^2^ test and the I^2^ index were used to assess the statistical heterogeneity. If there was no statistically significant heterogeneity (P>0.05 and I^2^<40%), a pooled effect was calculated with a fixed-effects model, which assumed that the observed differences among study results are due solely to chance^33^. If there was significant heterogeneity, the summary estimate and confidence/prediction interval would come from the random-effects model, which assumed that the different studies are estimating differently providing average intervention effect^33^.

If the study included more than two intervention arms, we reviewed only interventions that met the eligibility criteria or summarized at arm level. We only interpreted studies that were judged as low risk of overall bias in the main result section to avoid biases.

#### 2.1.8 Reporting bias assessment

If there were 5 or more studies in the meta-analysis, we assessed publication bias by graphical methods (funnel plots, which indicate the potential presence of reporting biases)^34^. In the plots, the x-axis represents the effect estimate, and the y-axis represents the standard error of the effect estimate. If there was publication bias, it would lead to an asymmetrical appearance of the plot^35^. Furthermore, the Egger test was used for reporting bias to evaluate the small study effect^36^.

#### 2.1.9 Additional analysis

If the heterogeneity was significant in the meta-analysis, the linear random-effect meta-regression was used to explore the trial-level covariates that contributed to the heterogeneity^37^. The candidate covariates were zinc dose, trial duration, and whether the trial specified the participant’s diabetes type (T2D, not specified types). The first two were treated as continuous variables and the third was a binary variable.

We illustrated the effect of interest if only one study reported a type of secondary outcome. If more than one study reported a type of secondary outcome, a meta-analysis was conducted regardless of the risk-of-bias. We also performed a sensitivity analysis which included all trials into meta-analysis regardless of risk assessment for comparing the results to evaluate the impact of bias.

### 2.2 two-sample Mendelian randomization

#### This two-sample MR study followed STROBE-MR Guideline^38^

#### 2.2.1 Study design and data sources

Sample sizes and GWAS data including both exposure and outcome was utilized from the MR-base. MR-base is the platform that thousands of GWAS summary datasets were updated from different research group. It contained the database of GWAS summary association statistics which structured into complete summary data for SNP-phenotype associations^39^.

We included all “zinc supplement” SNPs (Open GWAS ID: ukb-b-10567, ukb-b-13891, ukb-a-496) that meet the p-value threshold of <5x10^-8^ from the MR-base GWAS catalog. We also extracted the summary data (beta and SE) for the diabetes outcomes: fasting blood glucose (Open GWAS ID: ieu-b-114, GCST000568, ebi-a-GCST007858, ebi-a-GCST005186, ieu-b-113), HbA1C (Open GWAS ID: ieu-b-103, ieu-b-104), HOMA-IR (Open GWAS ID: ieu-b-118, ebi-a-GCST005179), and fasting insulin (Open GWAS ID: ebi-a-GCST005185, ebi-a-GCST000571, ebi-a-GCST007857, ieu-b-115, ieu-b-116). Proxy SNPs were included based on an LD of 0.001 and kb 10000 from the 1000 genomes European reference sample. Description of the methods used for these GWASs can be found in reference^40–42^.

For exposure data, the clumping genetic distance for LD was also set to 10000 kilobases. For outcome data, the minimum linkage disequilibrium (LD) value was set to 0.8 to prevent the non-random occurrence of genetic variants^43^. And, there was a threshold for minor allele frequency at <0.3, for ambiguities, allowing palindromic SNPs^44, 45^. We allowed MR-Base to automatically harmonize the exposure and outcome data, and attempt to align palindromic SNPs based off their minor allele frequency.

#### 2.2.2 Assumptions and assessment

There are three routine assumptions and two additional assumptions in two-sample MR.

1. Relevance: The genetic variants are associated with the exposure of interest.

✓ This assumption was assessed via F statistic for the gene-exposure relationship.
2. Independence: The genetic variants share no unmeasured cause with the outcome.

✓ This assumption was by ensuring that the GWASs have adequately controlled for plausible confounders of the gene-outcome association by adjusting for at least 10 principle components or using a linear mixed model.
3. Exclusion restriction: The genetic variants do not affect the outcome except through their potential effect on the exposure of interest.

✓ Pleiotropy can violate this assumption when genes influence two or more traits. Serval secondary analyses were used to detect and adjust the pleiotropy such as MR-Egger regression, weighted median, and weighted mode analysis.
4. The samples for exposure and outcome assessment are independent.

✓ We excluded the outcome summary data which had the same consortium as the exposure data to meet this assumption.
5. The samples for exposure and outcome assessment are from the same population.

✓ We chose the same outcome population category such as European with the exposure population, ensuring the fifth assumption.

#### 2.2.3 Statistical methods

We used the “TwoSampleMR” package in the R environment (version 4.1.0) to conduct the 2-sample MR analyses^39^. This package supported the harmonization process to ensure that both SNPs were coded from the same strand when SNPs were palindromic^44^. We used the inverse variance weighting (IVW) estimator to evaluate the causal inference. It is meta-analyzing Wald ratios which is the ratio of the SNP-outcome association/SNP-exposure association. Due to the difference in MR-base, the two outcomes were slightly changed compared to the review: fasting glucose and fasting insulin.

Cochran Q statistics can be used to assess evidence of heterogeneity in consideration of pleiotropy^46^. The statistically significant level was at the P<0.05. Steiger filtering checked the direction of causation between the exposure and outcome for each SNP^47^.

We therefore used the exposure of calcium supplement and outcome of hair color as the negative controls to detect the potential residual bias due to nutrients supplementation and population structure respectively. These were chosen because calcium supplementation is a type of nutritional supplement which is not thought to impact diabetes, and hair color is known to vary with population structure within the UK but is unlay to have any true causal association with zinc supplementation.

## 3. Results

### 3.1 Systematic review

#### 3.1.1 Study selection

A total of 15 studies, reporting on 14 trials, were identified for inclusion in the review (Fig. 1). One trial generated two studies (Parham 2008^48^ and Heidarian 2009^49^). From the four databases, 1557 citations were found, and 3 studies were identified by reference reading. 1531 citations were removed because of duplication or unfulfilling inclusion criteria with title or abstract. After the full-text screening of 29 studies, 14 studies were excluded because they did not meet the criteria. There were 11, 12, 7, and 8 studies measuring fasting blood glucose, HbA1C, HOMA-IR, and serum insulin levels respectively.

**Figure 1:**
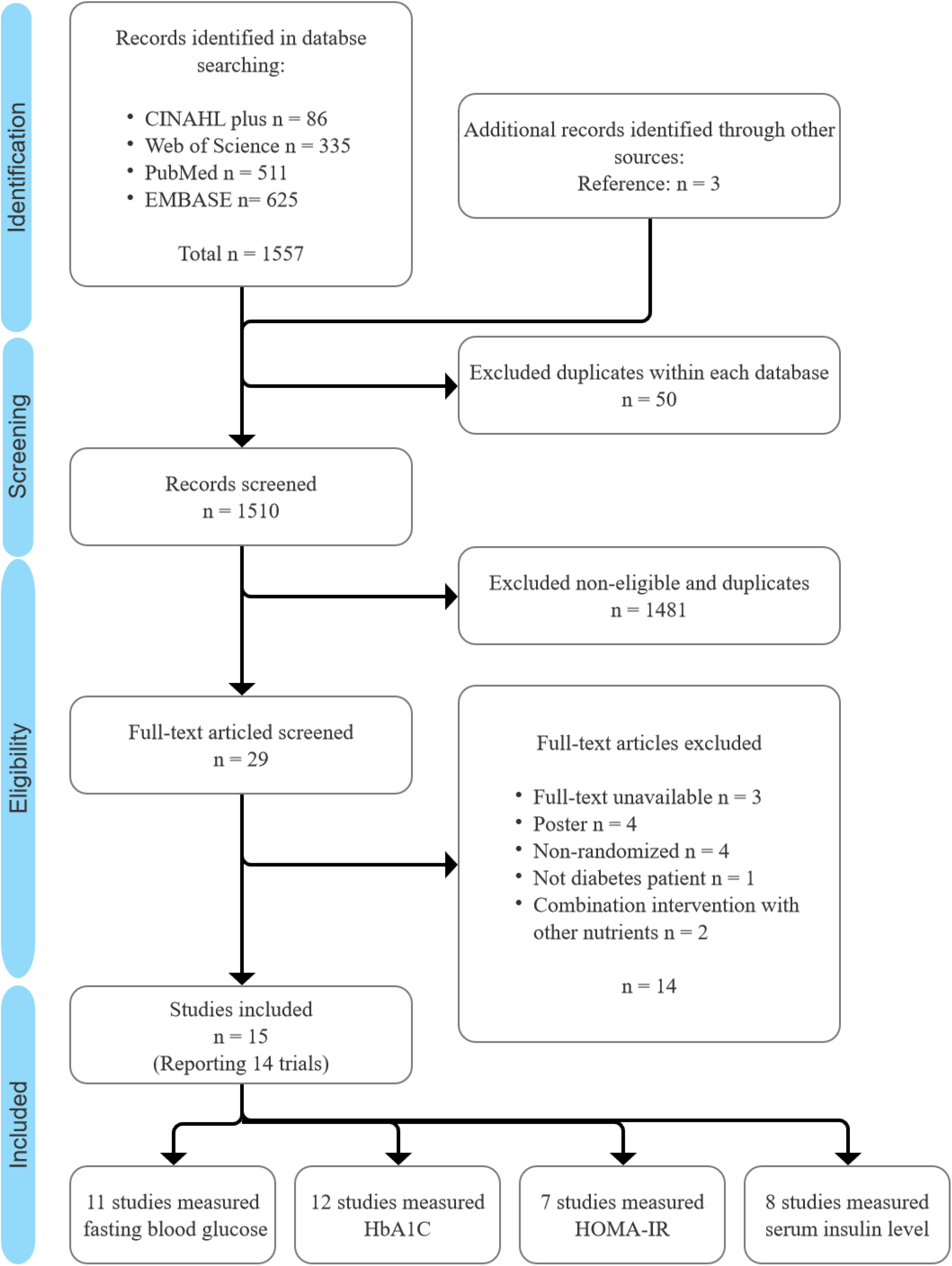
PRISMA flow diagram of information through the different phases of the systematic review

The percentage agreement was 96.32% and Gwet AC was 0.9619 between two reviewers, with strong evidence of significance (P<0.05), which indicated a very good inter-reviewer reliability for the selection process. No unpublished relevant studies were obtained.

#### 3.1.2 Study characteristics

There were 13 trials in parallel design and one in crossover design (Parham 2008^48^ and Heidarian 2009^49^). Details of each trial are presented in table 1. A total of 897 participants were randomly allocated, and all trials had a relatively small sample size. The time of publication ranged from 2003 to 2021. Most trials had claimed that there was no significant imbalance (n=7), or they used matched methods (n=2) for age and sex. One trial clearly demonstrated that their participants were adolescents, while the other’s average age was above 45-year-old. Four trials did not specify the type of diabetes in the inclusion criteria and patients had complications: diabetic foot ulcer^50^, diabetic hemodialysis with Zn-deficiency^51^, diabetic retinopathy^52^, and b-thalassemia major complicated^53^.

**Table 1:** Summary of included studies evaluating the effect of zinc intervention

| Study ID | RCT design | Country | Total N. randomized | Duration (month) | Gender, M/F | Age mean (control vs intervention) or age inclusion range | Formulation/elemental zinc dose, mg/day | Health status |
| --- | --- | --- | --- | --- | --- | --- | --- | --- |
| Afkhami - Ardekani 2008<sup>54</sup> | Parallel | Iran | 40 | 1.5 | 16/24 | 52.67 ± 8.60 | zinc sulfate/149.82 | T2D |
| Asghari 2019<sup>55</sup> | Parallel | Iran | 64 | 3 | 32/28 | 45.5 ± 5.4 vs 46.2 ± 5.3 | zinc gluconate/4.29 | T2D |
| Burki 2017<sup>51</sup> | Parallel | Pakistan | 101 | 3 | NA | NA | zinc sulphate/4.54<sup>a</sup> | T2D |
| Gunasekara 2011<sup>53*</sup> | Parallel | Sri Lanka | 96 | 4 | 33/63 | 51.2 ± 6.0 vs 54.1 ± 6.0 | elemental zinc/22<sup>b</sup> | T2D |
| Hosseini 2021<sup>48</sup> | Parallel | Iran | 46 | 2 | 25/21 | 54.1 ± 5.4 vs 55.6 ± 7.3 | elemental zinc/50 | Diabetic hemodialysis patients with Zn-deficiency |
| Khan 2013<sup>52</sup> | Parallel | India | 54 | 3 | NA | 56.0 ± 0.86 vs 56.3 ± 6.6 | zinc sulphate/11.35 | T2D |
| Matter 2020<sup>50</sup> | Parallel | Egypt | 80 | 3 | 18/32 | 16.1 ± 1.4 vs 16.4 ± 1.4 | zinc gluconate/7.72 | Diabetes with b-thalassemia major complicated |
| Momen-Heravi 2017<sup>47</sup> | Parallel | Iran | 60 | 3 | 42/18 | 60.0 ± 10.0 vs 58.3 ± 8.6 | elemental zinc/50 | Diabetes with diabetic foot ulcer |
| Naghizadeh 2018<sup>49</sup> | Parallel | Iran | 50 | 3 | 20/25 | 58.0 ± 5.9 vs 57.4 ± 5.8 | zinc gluconate/4.29 | Patients with diabetic retinopathy |
| Nazem 2019<sup>56</sup> | Parallel | Iran | 70 | 2 | 19/16 | 54.3 ± 7.2 vs 53.3 ± 7.4 | zinc gluconate/7.15 | T2D |
| Parham 2008<sup>45</sup>,<br>Heidarian 2009<sup>46</sup> | Crossover | Iran | 50 | 2.5 | 24/18 | 52.0 ± 9.3 / 54.5 ± 9.2 | elemental zinc/30 | T2D |
| Pérez 2018<sup>57</sup> | Parallel | Chile | 80 | 12 | 13/15 | 30 - 65 | elemental zinc/6.81 | T2D with further selection criteria |
| Roussel 2003<sup>58</sup> | Parallel | Tunis | 56 | 6 | NA | 55.5 ± 1.4 vs 51.5 ± 1.6 | zinc gluconate/4.29 | T2D |
| Witwit 2021<sup>59</sup> | Parallel | Iraq | 50 | 1.5 | NA | 25 - 60 | zinc sulphate/11.35 | T2D |

\*: It was a three-arm study and we only extracted zinc and its control arm.
a: Oral hypoglycemic arm and zinc plus oral hypoglycemic arm.
b: multivitamin/mineral arm and zinc plus multivitamin/mineral arm.

The duration of zinc supplementation ranged from 1.5 to 12 months, with a mean and median duration of 3.53 months and 3 months. Three trials contained basic supplements care besides the zinc administration and placebo: oral hypoglycemic agent (in Burki 2017^54^ and Khan 2013^55^) and multivitamin/mineral (in Gunasekara 2011^56^). Each dose of zinc gluconate and zinc sulfate was multiplied by 0.143 or 0.227 to obtain the appropriate dose of elemental zinc^21^. The mean dose of elemental zinc included in these interventions was 25.83 mg/d (range: 4.29–149.82 mg/d; median: 9.25 mg/d).

#### 3.1.3 Results of individual studies

Nine trials measured post-trial means with SE of fasting blood glucose as outcomes. Seven of the nine trials were at low risk of bias. Eight trials including the crossover trial measured post-trial HbA1C as an outcome. Six of the eight trials were at low risk of bias. Six trials with low risk-of-bias had eligible measured post-trial HOMA-IR scores. Six trials with low risk-of-bias had eligible measured post-trial serum insulin as an outcome. Notably, we standardized the measurement of serum insulin on a single scale through Hedges’ g score and presented the SMD.

#### 3.1.4 Synthesis of results

According to the I^2^ and p-value of the Cochran-Q χ2 test, the random-effects model was used for meta-analysis for all outcomes. Results were presented in figure 2

**Figure 2:**
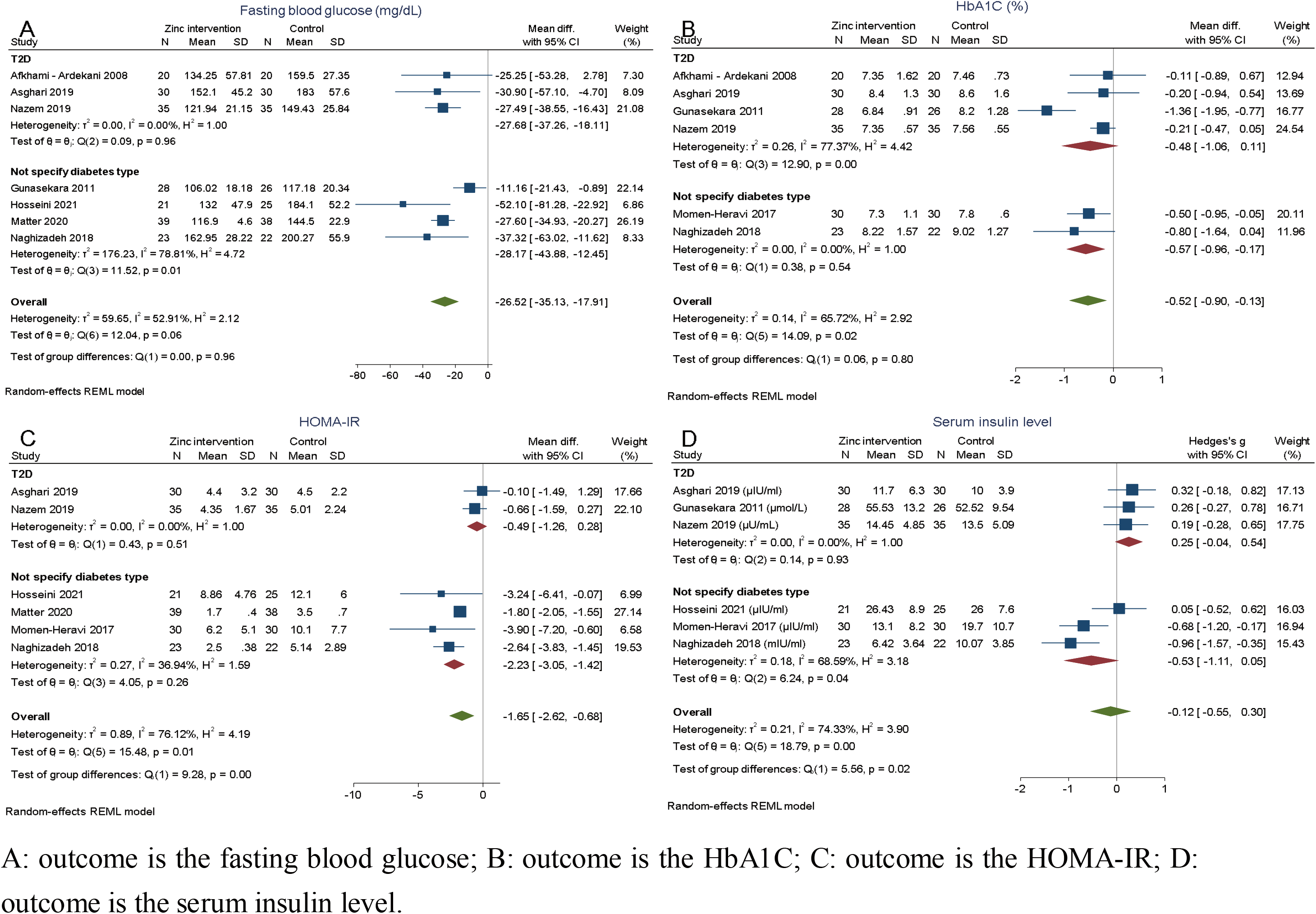
Forest plot of the standardized mean difference of each outcome insulin level between zinc intervention and control arms for low risk-of-bias trials

##### Fasting blood glucose

Among the T2D patients, the pooled mean difference was -27.68 mg/dL (95%CI: -37.26, -18.11), comparing zinc intervention to the control arm (Fig. 2A). Among the participants without specified diabetes type, the pooled mean difference was -28.17 mg/dL (95%CI: -43.88, -12.45). Both stratified analyses showed strong evidence of effect and the two types of trials were not significantly different (Test of group difference P=0.96). The overall pooled mean difference showed that the zinc intervention had a significantly lower level of fasting blood glucose at the end of the trial, compared to the control arm (MD: -26.52, 95%CI: -35.11, -17.91).

##### Hemoglobin A1C

T2D patients have a decrease (MD: 0.48, 95%CI: -1.06, 0.11) in HbA1C, but this difference was not significant between arms (Fig. 2B). Among the participants without specified type, the pooled mean difference was -0.57% (95%CI: -0.96, -0.17) in HbA1C, compared to zinc intervention and control arm. There was no significant difference between those two types of trials (Test of group difference P=0.80). Overall, there was a significant reduction in post-trial HbA1C percentage between zinc intervention and control at end of the trial (MD: -0.52, 95%CI: -0.90, -0.13).

##### HOMA-IR

The pooled mean difference was -0.49 (95%CI: -1.26, 0.28) in T2D patients and -2.23 (95%CI: -3.05, -1.42) among patients with unspecified type in the post-trial HOMA-IR (Fig. 2C). There was a significant difference detected between those two types of trials (Test of group difference P<0.01). The pooled result shows that the zinc intervention arm had a 1.65 lower (95%CI: -0.90, -0.13) HOMA-IR score than the control arm with strong evidence.

##### Serum insulin level

For serum insulin level (Fig. 2D), the SMD was 0.25 (95%CI: -0.04, 0.54) in T2D patients and -0.53 (95%CI: -1.11, 0.05) among the participants without specified diabetes type, with only weak evidence. Overall, the pooled result shows that there was no significant standardized mean difference in serum insulin level between zinc intervention and control arms at end of the trial (SMD: -0.12, 95%CI: -0.55, 0.30).

The results of score change are presented in supplementary material 4. Some secondary outcomes were found after full-text reading, and their results in post-hoc analysis are presented in supplementary material 5.

#### 3.1.5 Reporting bias assessment

We observed a slight asymmetry in the funnel plot of the fasting blood glucose in figure 3A. One study fell out of the 95% confidence limits. The p-value of the Egger test for small-study effects was 0.1649 (>0.05), which meant there was no strong reporting bias.

**Figure 3:**
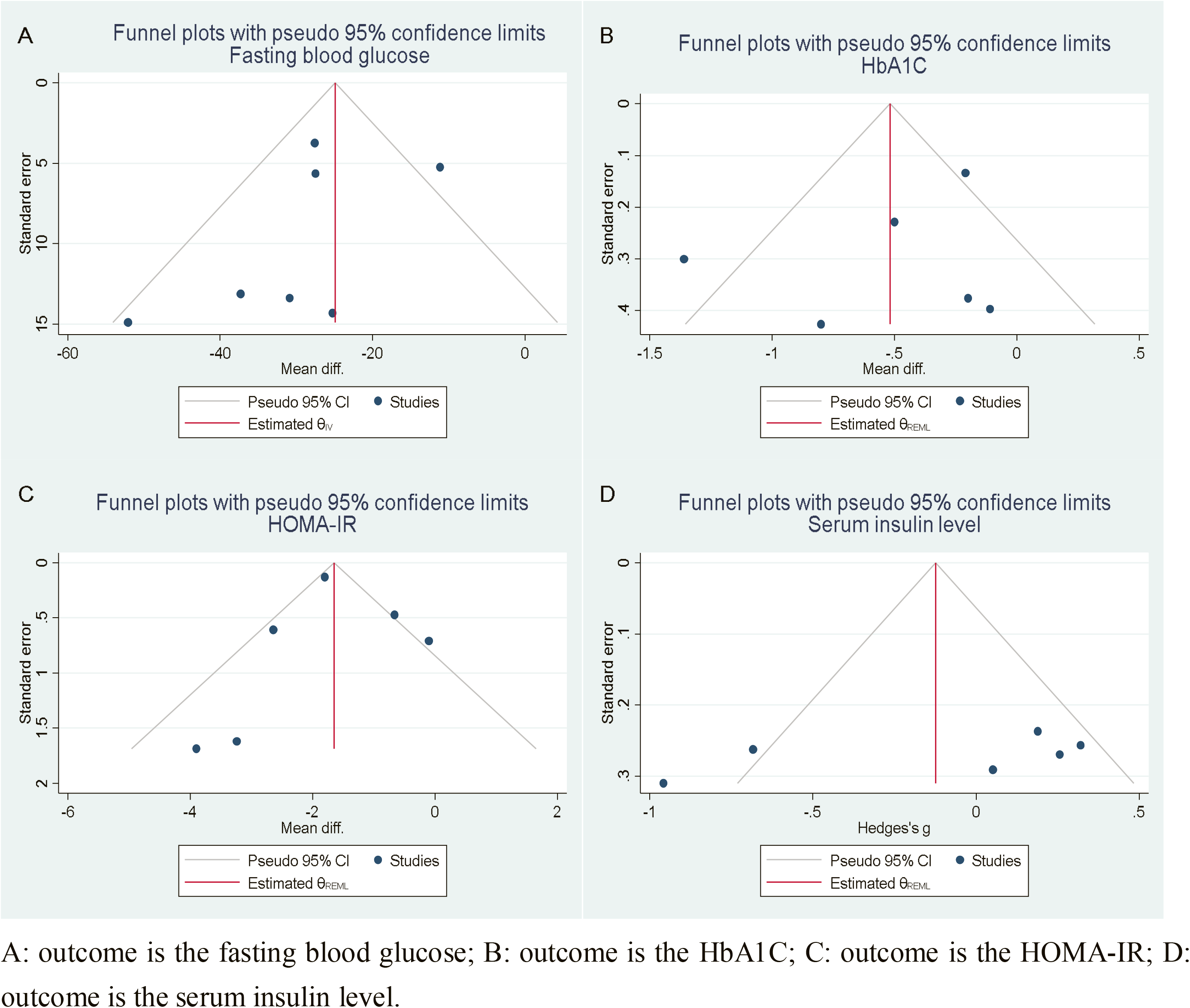
Funnel plot with pseudo 95% confidence limits for the standardized mean difference of each outcome at post-trial

Visual assessment of the funnel plots of HbA1C (Fig. 3B), HOMA-IR (Fig. 3C), and serum insulin level (Fig. 3D) suggested that there was no asymmetry. Further, the p-value of the Egger test for small-study effects was 0.7341, 0.2697, and 0.1514 for HbA1c, HOMA-IR, and serum insulin level, respectively, which rejected the null hypothesis of no asymmetry and suggested no significant reporting bias exist.

#### 3.1.6 Additional analysis: meta-regression

Second, through meta-regression, we found evidence to support a linear association of fasting blood glucose (regression coefficient: 8.42, 95%CI: 0.52, 16.33) and HbA1C (regression coefficient: -0.46, 95%CI: -0.73, -0.19) with trial duration, which was the significant explanatory variable on heterogeneity. Whether specified diabetes types significantly explained the heterogeneity for the outcome of HOMA-IR (regression coefficient: -1.68, 95%CI: -2.84, -0.51) and serum insulin level (regression coefficient: -0.78, 95%CI: -1.31, -0.25) in the meta-analyses. We shall be conscious about the results because none of the outcomes had more than 10 trials. These results are presented in Table 2.

**Table 2:** Meta-regression of potential explanatory variables on heterogeneity in each outcomès meta-analysis

| Outcome | N. of study | Dose of zinc intervention |  | Duration of trial |  | Diabetes type specified |  |
| --- | --- | --- | --- | --- | --- | --- | --- |
|  |  | Mean (mg) | Coefficient (95%CI) | Mean (mo.) | Coefficient (95%CI) | N. of T2D specified | Coefficient (95%CI) |
| Fasting blood glucose | 7 | 34.75 | -0.01 (-0.24, 0.23) | 2.64 | 8.42 (0.52, 16.33)* | 3 | 1.31 (-18.43, 21.05) |
| HbA1C | 6 | 39.59 | 0.00 (-0.01, 0.01) | 2.75 | -0.46 (-0.73, -0.19)* | 4 | -0.13 (-1.06, 0.77) |
| HOMA-IR | 6 | 20.24 | -0.05 (-0.11, 0.01) | 2.67 | -0.46 (-2.85, 1.93) | 2 | -1.68 (-2.84, -0.51)* |
| Serum insulin level | 6 | 22.96 | -0.00 (-0.03, 0.02) | 2.83 | -0.04 (-0.73, 0.65) | 3 | -0.78 (-1.31, -0.25)* |

### 3.2 two-sample Mendelian randomization

#### 3.2.1 Description of exposure-SNP summary

Three SNPs were extracted out, which represented zinc supplement: rs6756297, rs4861163, and rs10822145, and their GWAS all were conducted from the UK biobank (Tab. 3). Their number of participants for GWAS was 461384.

**Table 3:** Description of the exposure-SNP association

| SNP | Exposure: Zinc supplement |  |  |  |  |
| --- | --- | --- | --- | --- | --- |
|  | Sample size | Beta | SE | Population | Consortium |
| rs4861163 | 461384 | 0.006 | 0.001 | European | UK Biobank |
| rs10822145 | 461384 | 0.002 | 0.000 | European | UK Biobank |
| rs6756297 | 461384 | -0.003 | 0.000 | European | UK Biobank |

#### 3.2.2 Description of outcome-SNP summary

The summary statistics of each outcome with each SNP were reported in table 4. Since the exposure data was from the UK biobank, we chose the MAGIC consortium with the European population for each outcome.

**Table 4:**
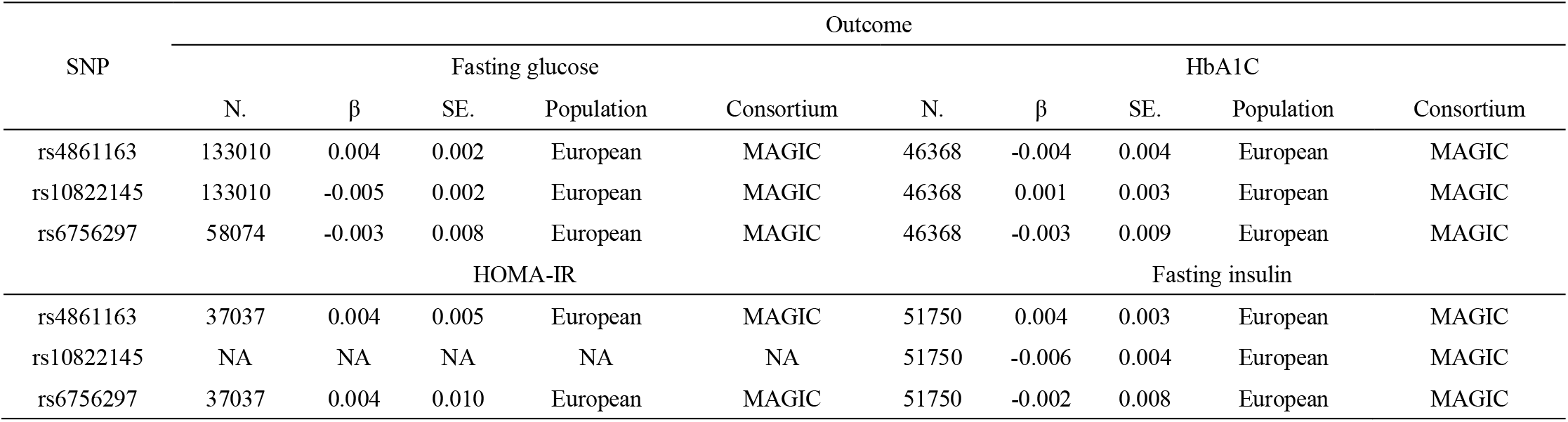
Description of the outcome-SNP association

#### 3.2.3 Results of two-sample MR

The results including the heterogeneity test were presented in figure 4. Zinc supplement with 2 SNPs led to signification decrease of fasting glucose (coefficient: -2.04, 95%CI: -3.26, -0.83). The unit of fasting glucose was mmol/L^63^ (-2.04 mmol/L = -37.09 mg/dL). While there was no strong evidence supporting the relationship of zinc supplements with HbA1C, HOMA-IR, and insulin level. The p-value for heterogeneity of fasting glucose, HbA1C, HOMA-IR, and insulin level were 0.29, 0.66, 0.37, and 0.47. Neither of these results had significant heterogeneity, which suggested no pleiotropy with the null hypothesis.

**Figure 4:**
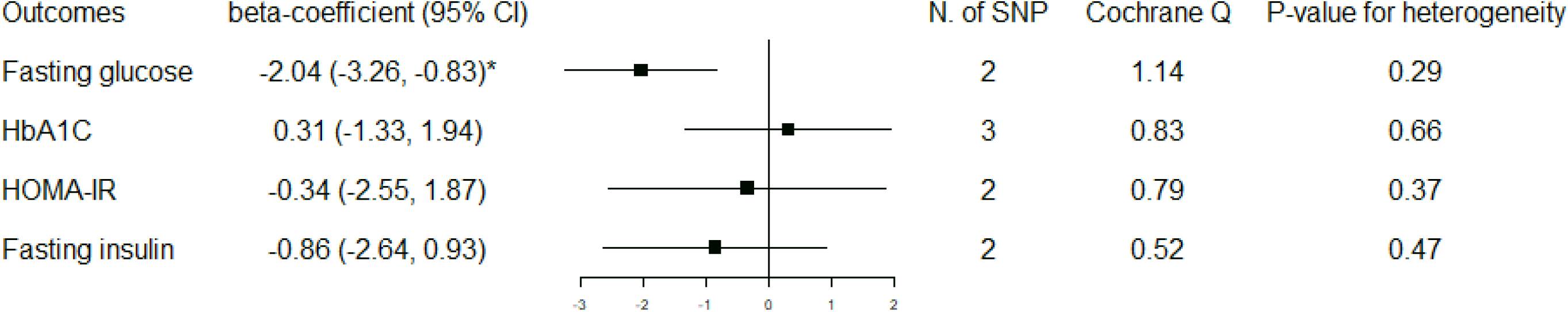
two-sample MR results with IVW coefficient

## 4. Discussion

### Principal findings

This systematic review, including 14 RCTs between 2003 and 2021, reported how single zinc supplements impacted glycemic control in type-2 diabetes. In meta-analyses with random-effect models, zinc supplements were shown to lead to a significant reduction in the level of post-trial fasting blood glucose, HbA1C, and HOMA-IR, but not in the serum insulin level. These results had moderate certainty of evidence. Trial duration contributed to the heterogeneity in the results of fasting blood glucose and HbA1C and diabetes type specified contributed to the heterogeneity in HOMA-IR and serum insulin level. In the two-sample MR analysis, with 2 SNPs, zinc supplement also significantly associated with a lower level of fasting glucose. The difference in effect sizes between review and two-sample MR were small (-26.52 vs -37.09, overlapping in CI).

### Previous literature

The protective power of zinc supplements was demonstrated in the previous literature across different populations and diabetes types. In a cohort study encompassing 14,140 Japanese, the dietary intake of zinc was associated with a lower odds ratio of type-2 diabetes mellitus among the younger (age 40-55 years)^64^. The result of lower T2D risk was also consistent with an Australian women’s longitudinal cohort^65^. Furthermore, de Carvalho and colleagues found a negative correlation between %HbA1c and plasma zinc levels, and women with deficient zinc levels had higher scores of HOMA-IR and C peptide values in a systematic review^66^. Besides, a study with two cohorts observed an 11% decrease in the risk of gestational hyperglycemia by every 1 mg/d zinc intake in women, which might be used to prevent gestational diabetes^67^.

In 2019, Wang et. al indicated that zinc supplementation could significantly reduce key glycemic indicators in a systematic review of RCT that included all diabetes types. This review involved 1700 participants and found a larger net change among fasting glucose, 2-h postprandial glucose, HbA1C, and HOMA-IR in zinc intervention arm^20^. Zinc supplements had a larger effect on fasting glucose in diabetes patients than people at high risk and a preventative effect among prediabetes^20^. These findings were consistent with our results on zinc’s favorable effect on glycemic control and the magnitude of HbA1C’s MD was similar to ours. But they did not specify the diabetes type in their population included. A systematic review of RCT published in early 2021, specified zinc as a single factor of intervention. and found that low-dose and short-duration zinc supplements also showed significant improvements in some T2D indicators, compared with the high-dose and long-duration^21^. The specific effect also supported our result in meta-regression about the heterogeneity contribution of the trial duration, which plays a critical role in the zinc supplement’s effect. However, the true effect of zinc might be masked by the multi-supplement intervention or non-specified diabetes type with differential biological mechanisms.

There were not many MR studies that directly linked zinc as exposure to diabetes. An MR study in 2019 did not find any significant causal relationship between zinc level in blood with 2 SNPs and odds of T2D^68^. The difference with our results may be due to the supplementary effect of zinc.

### Strength

To our knowledge, this study is the first to use mixed methods by the systematic review of RCT and MR to triangulate evidence of causal inference in the field of nutritional supplementation. This systematic review included the newest, until 2021, literature with scientific rigor and delivered a clear statement of the zinc supplement’s protection on type-2 diabetes at the individual level. Our result of improving glycemic control was consistent with previous literature. Not only that, we narrowed the intervention into a single supplement to emphasize the effect of zinc with less concern of contamination bias. Moreover, on the basis of the review, the application of two-sample Mendelian randomization minimized residual confounding or reverse causality due to imperfect randomization or short duration and gave a more comprehensive lifelong evaluation. MR assumed that the effect of instrumental SNPs was only from the exposure of interest to the outcome, independent of other factors. Compared with one-sample MR, two-sample increased statistical power with larger population from multiple GWAS consortia^69^.

### Limitation of included studies, systematic review

The majority of the studies had relatively small groups in each arm (around 30 participants). Although this may limit the power and confounder adjustment, the method of meta-analysis could combine them to get stronger results. Moreover, the trials with at least some concerns of bias were not included in the main interpretation. In the sensitivity analysis of all trials, the effect estimate of sensitivity results was similar to the main, which reduced our concern (supplementary material 6). Suspicious publication bias of fasting blood glucose perhaps came from the small number of trials included. Also, there may exist the problem of generalizability that most of the trials were in Asia, which may induce the problem of extrapolation. A possible explanation is that researchers from high-income countries prefer to perform trials with co-supplement and target a wider population of all types of diabetes. Thus, further RCT with larger sample sizes in a variety of geographic regions would be preferred.

### Limitation of review processes, systematic review

In the review process, the study was limited in that only one reviewer screened every paper and extracted information. The high percentage agreement and Gwet AC in the random set reduced our concerns. Second, we found some degree of heterogeneity in each outcome analysis. Due to the limited number of trials, the meta-regression was not strong enough to detect explanatory variables. We may expand our criteria to include more trials in the situation of not affecting the directness.

### Limitation of two-sample MR

The MR part was limited to the low number of SNPs included. This meant that sensitivity analysis like MR-Egger for exclusion restriction assumptions, and leave-one-out could not be used. MR studies are often low powered, the small number of SNPs will have additionally reduced power of the MR analyses. Although there is a potential bias from the effect of supplementary, the negative control using calcium supplementation implied that this is unlikely to have produced a substantive bias (supplementary material 7). The two-sample MR results of negative control: hair color suggested no significant association between zinc supplementary and hair color (supplementary material 8), which reduced the threat from bias due to residual population structure. Third, pooled analysis of individual studies made us not access to the individual patient data to specify the T2D population.

### Biological plausibility

Zinc’s antioxidant properties are the biologically plausible connection to T2D. Oxidative stress reflected an imbalance between the production of reactive oxygen species (ROS) and antioxidant defenses, and excess ROS could cause lipid peroxidation, leading to the lesion of cell membranes and lipoproteins, and ultimately damage insulin secretion to increase resistance through signaling pathway within the β-cells of pancreatic islets^70, 71^. Plasma advance oxidation protein products generated during oxidative stress were indicated as a biomarker of endothelial dysfunction as early events for T2D diabetes^72^. As a catalytic role for zinc superoxide dismutase, zinc involves the conversion of superoxide radicals to molecular oxygen and hydrogen peroxide through the antagonism, which could reduce the reactive oxygen species (ROS) toxicity^73, 74^. For chronic effects, zinc is also an inducer of metallothionein in multiple organs^74^. The metallothionein immunoreactivity levels were higher in the tubular areas of the zinc-supplemented group, suggesting zinc’s stimulation effect to regulate the oxidative stress in rat^75^. The sulfhydryl stabilization by zinc protection was another mechanism against the oxidative damange^76^.

Many studies have investigated the metabolism of zinc in the pathogenesis of diabetes mellitus. In the spontaneously diabetic mice, researchers observed the `in vitro` insulinomimetic activity, hypoglycemic effect in glucose, and insulin resistance attenuation by administration of Zn complexes^77^. In the population study, diabetic patients had a higher level of 8-hydroxy-2-deoxyguanosine, oxidative damage to DNA, and a lower level of zinc, which suggested more serious oxidative lesion^78^. By analogy, people with higher dietary antioxidant capacity (richer antioxidants in diet) associated with a lower risk of T2D and a lower score of HOMA-IR across sexes in the Rotterdam study^79^. Antioxidants become therapeutic options to manage diabetes and its serious complications such as Metformin, which reduce the production of ROS and increase insulin sensitivity^80^. So that, zinc administration has huge potential to be a first-line intervention treatment for T2D due to its antioxidant mechanism.

### Implications

Optimal ways to utilize the zinc supplement intervention in real-world health practices need more concrete explorations, in particular in the context of disrupted routine care due to the COVID pandemic. According to a rapid WHO assessment, nearly 50% of diabetes and diabetic complications management services were partially or completely disrupted, which was an essential service in most of the countries, especially among LMICs ^81^. Timely nutritional interventions are essential for successful and sustainable diabetes care to overcome the negative effect from the implication of strict infection prevention and control^82^. Self-care practice should be encouraged during this extreme health workforce shortage^83^. Immediate implementation of zinc as an efficient tool or complement is needed for patients who struggle with limited medical resources and primary care systems which are considerably disrupted.

## 5. Conclusion

The study found that a single zinc supplement was strongly associated with a lower level of glycemic indicators in type-2 diabetes, suggesting a protective effect. The systematic review and two-sample Mendelian randomization supported a causal association, but need to be further validated. We advocate the use of zinc supplements as an efficient nutritional intervention to support routine diabetic care.

## Data Availability

All data produced are available online at MR Base and academic databases.

