## Supplementary material 1-8 for "Triangulating evidence for the causal impact of single-dose zinc supplement on glycemic control for type-2 diabetes"

### Content of supplementary material

Supplementary material 1: Searching strategy for each database

Supplementary material 2: Risk-of-bias assessment

Supplementary material 3: Certainty assessment

Supplementary material 4: Forest plot of the standardized mean difference of each outcome`s change score between zinc intervention and control arms for low risk-of-bias trials

Supplementary material 5: Post-hoc analysis between zinc intervention and other outcomes

Supplementary material 6: Forest plot of the mean difference or standardized mean difference of post-trial outcomes between zinc intervention and control arm for all trials.

Supplementary material 7: Association between calcium supplementary and diabetes outcome in two-sample MR

Supplementary material 8: Association between zinc supplementary and hair color in two-sample MR

Reference

**Supplementary material 1: Searching strategy for each database**

**PubMed:**

((zinc[MeSH Terms]) OR (zinc[Title/Abstract])) AND ((diabet*[Title/Abstract]) OR (DM[Title/Abstract]) OR (T2M[Title/Abstract]) OR (Diabetes Mellitus[MeSH Terms])) AND ((“blood sugar”[Title/Abstract]) OR (“blood glucose”[Title/Abstract]) OR (glycemic[Title/Abstract]) OR (hyperglycemi*[Title/Abstract]) OR (blood glucose[MeSH Terms]) OR (HbA1c[Title/Abstract]) OR (glycemic control[MeSH Terms])) AND ((control* [Title/Abstract]) OR (blind* [Title/Abstract]) OR (random*[Title/Abstract]) OR (trial*[Title/Abstract]) OR (RCT[Title/Abstract]) OR (placebo*[Title/Abstract]) OR (randomized controlled trial[MeSH Terms]))

**CINAHL Plus:**

(AB zinc OR MH zinc) AND (AB diabet* OR AB DM OR AB T2M OR MH Diabetes Mellitus) AND (AB “blood sugar” OR AB “blood glucose” OR AB glycemic OR AB hyperglycemi* OR AB HbA1c OR MH glycemic control OR MH blood glucose) AND (AB control* OR AB blind* OR AB random* OR AB trial* OR AB RCT* OR AB placebo* OR MH randomized controlled trial)

**EMBASE:**

1 zinc.ab,ti.

2 "diabet*".ab,ti.

3 DM.ab,ti.

4 T2M.ab,ti.

5 blood glucose.ab,ti.

6 blood sugar.ab,ti.

7 glycemic.ab,ti.

8 "hyperglycemi*".ab,ti.

9 HbA1c

10 "control*".ab,ti.

11 "blind*".ab,ti.

12 "random*".ab,ti.

13 "trial*".ab,ti.

14 "RCT*".ab,ti.

15 "placebo*".ab,ti.

16 2 or 3 or 4

17 5 or 6 or 7 or 8 OR 9

18 10 or 11 or 12 or 13 or 14 or 15

19 1 and 16 and 17 and 18

**Web of Science:**

AB=(zinc AND (diabet* OR DM OR T2M ) AND ( “blood sugar” OR “blood glucose” OR glycemic OR hyperglycemi* OR HbA1c) AND (control* OR random* OR trial* OR RCT* OR placebo*) )

**Supplementary material 2: Risk-of-bias assessment**

**Assessment rule**

The Cochrane Risk of Bias tool (ROB 2) was followed as a framework to assess the risk of bias within each study by one unblinded reviewer. It is structured into five domains covered all types of bias that can affect the results of randomized trials:

1. bias arising from the randomization process.
2. bias due to deviations from intended interventions.
3. bias due to missing outcome data.
4. bias in the measurement of the outcome.
5. bias in the selection of the reported result.

The ROB 2 algorithm maps the answer of the signaling question (“no information”, “yes”, “probably yes”, “no”, or “probably no”) to a proposed judgment in each domain and then leads to an overall judgment[1]. The level of risk-of-bias is low risk, some concerns, and high risk. The Crossover RCT had an additional domain about carryover and period effect[1]

**Assessment results**

Burki 2017[2] trials raised some concern about the bias arising from the randomization process. Because they lacked information about baseline imbalance. Pérez 2018[3] and Khan 2013[4] had high risk, and Parham 2008[5]/Heidarian 2009[6] had some concern on the bias due to missing outcome data. We treated Khan 2013[4] at high risk because some of the participants in the follow-up were excluded from starting insulin use. Pérez 2018[3] was also considered as high risk for further selection criteria for the participant after allocation, which was highly correlated with the effect of interest (HbA1c between 5.7 and 8%). Parham 2008[5]/Heidarian 2009[6] had a small group of participants lost due to insulin using, but the crossover design reduced its effect to some extent because the loss was equal in intervention and control arm by self-comparison. Moreover, Roussel 2003[7] demonstrated some concern for bias in the selection of the reported results. Because they did not report the post-trial means for primary outcome outcomes but stated the relation between zinc intervention and outcomes.

All trials did not have the situation in which the zinc administration was carried in the control arm due to some purpose, so the bias risk due to intended intervention deviations was low. All trials used standard and appropriate laboratory assays to measure the outcome, reducing the bias in measurement. Parham 2008[5]/Heidarian 2009[6] had similar participants in two sequences, enough washout period and relatively short overall duration, which the risk arising from carryout and crossover effect was low.

In summary, five trials had at least "some concern" level of risk-of-bias. The overall and details of judgment are presented in Figure S1 and Table S1.

Figure S1: Risk of bias summary figure illustrating judgment about each risk of bias item for each included study, based on the revised Cochrane ROB2

| | Arising from the randomization process | Due to deviations from intended interventions | Due to missing outcome data | In measurement of the outcome | In selection of the reported results | Arising from carry and crossover effect (Only for crossover design) | Overall |
|---|---|---|---|---|---|---|---|
| Afkhami - Ardekani 2008 | Low | Low | Low | Low | Low | NA | Low |
| Asghari 2019 | Low | Low | Low | Low | Low | NA | Low |
| Burki 2017 | Some concern | Low | Low | Low | Low | NA | Some concern |
| Gunasekara 2011 | Low | Low | Low | Low | Low | NA | Low |
| Hosseini 2021 | Low | Low | Low | Low | Low | NA | Low |
| Khan 2013 | Low | Low | High | Low | Low | NA | High |
| Matter 2020 | Low | Low | Low | Low | Low | NA | Low |
| Momen-Heravi 2017 | Low | Low | Low | Low | Low | NA | Low |
| Naghizadeh 2018 | Low | Low | Low | Low | Low | NA | Low |
| Nazem 2019 | Low | Low | Low | Low | Low | NA | Low |
| Parham 2008, Heidarian 2009 | Low | Low | Some concern | Low | Low | Low | Some concern |
| Pérez 2018 | Low | Low | High | Low | Low | NA | High |
| Roussel 2003 | Low | Low | Low | Low | Some concern | NA | Some concern |
| Witwit 2021 | Low | Low | Low | Low | Low | NA | Low |

Table S1: Judgment and supporting evidence of risk-of-bias in each study.

| Study ID | Support or Judgement for risk of bias | | | | | | |
|---|---|---|---|---|---|---|---|
| | Randomization process | Deviations from intended interventions | Missing outcome data | Outcome measurement | Selection of the reported results | Carry and crossover effect | Overall |
| Afkhami - Ardekani 2008 | Randomized process and no baseline imbalance (Low) | No deviation found (Low) | Outcome data on all participants (Low) | Clear lab assay indicates (Low) | Reported eligible data (Low) | NA | low risk in all domains (Low) |
| Asghari 2019 | Randomized process and no baseline imbalance (Low) | No deviation found (Low) | Outcome data on all participants (Low) | Clear lab assay indicates (Low) | Reported eligible data (Low) | NA | low risk in all domains (Low) |
| Burki 2017 | Randomized process and unclear baseline (some concern) | No deviation found (Low) | Outcome data on all participants (Low) | Clear lab assay indicates (Low) | Reported eligible data (Low) | NA | Concern in some domain (Some concern) |
| Gunasekara 2011 | Randomized process and no baseline imbalance (Low) | No deviation found (Low) | Outcome data on all participants (Low) | Clear lab assay indicates (Low) | Reported eligible data (Low) | NA | low risk in all domains (Low) |
| Hosseini 2021 | Randomized process and no baseline imbalance (Low) | No deviation found (Low) | Outcome data on all participants (Low) | Clear lab assay indicates (Low) | Reported eligible data (Low) | NA | low risk in all domains (Low) |
| Khan 2013 | Randomized process and no baseline imbalance (Low) | No deviation found (Low) | Lost participants due to start using insulin (High) | Clear lab assay indicates (Low) | Reported eligible data (Low) | NA | High risk in some domain (High) |
| Matter 2020 | Randomized process and no baseline imbalance (Low) | No deviation found (Low) | Outcome data on all participants (Low) | Clear lab assay indicates (Low) | Reported eligible data (Low) | NA | low risk in all domains (Low) |

| | | | | | | | |
|---|---|---|---|---|---|---|---|
| Momen-Heravi 2017 | Randomized process and no baseline imbalance (Low) | No deviation found (Low) | Outcome data on all participants (Low) | Clear lab assay indicates (Low) | Reported eligible data (Low) | NA | low risk in all domains (Low) |
| Naghizadeh 2018 | Randomized process and no baseline imbalance (Low) | No deviation found (Low) | Outcome data on all participants (Low) | Clear lab assay indicates (Low) | Reported eligible data (Low) | NA | low risk in all domains (Low) |
| Nazem 2019 | Randomized process and no baseline imbalance (Low) | No deviation found (Low) | Outcome data on all participants (Low) | Clear lab assay indicates (Low) | Reported eligible data (Low) | NA | low risk in all domains (Low) |
| Parham 2008, Heidarian 2009 | Randomized process and no baseline imbalance (Low) | No deviation found (Low) | Lost participants due to start using insulin in both sequences (some concern) | Clear lab assay indicates (Low) | Reported eligible data (Low) | No carry and crossover effect apparent (Low) | Concern in some domain (Some concern) |
| Pérez 2018 | Randomized process and no baseline imbalance (Low) | No deviation found (Low) | Further selection after randomization and criteria correlated with T2D (High) | Clear lab assay indicates (Low) | Reported eligible data (Low) | NA | High risk in some domain (High) |
| Roussel 2003 | Randomized process and no baseline imbalance (Low) | No deviation found (Low) | Outcome data on all participants (Low) | Clear lab assay indicates (Low) | Only report association with zinc but no post-trial mean (some concern) | NA | Concern in some domain (Some concern) |
| Witwit 2021 | Randomized process and no baseline imbalance (Low) | No deviation found (Low) | Outcome data on all participants (Low) | Clear lab assay indicates (Low) | Reported eligible data (Low) | NA | low risk in all domains (Low) |

**Supplementary material 3: Certainty assessment**

**Judgement rule**

Grading of Recommendations, Assessment, Development, and Evaluation (GRADE) summarizes confidence and rates the certainty of the evidence for the meta-analysis, which takes five considerations: risk of bias, consistency of effect, imprecision, indirectness, and publication bias[8]. Nevertheless, the GRADE assessment contained a considerable amount of subjectivity in each decision by necessity[9]. There were four levels of the certainty grade: high, moderate, low, and very low.

The consideration of risk-of-bias was regarded as high certainty because we only interpret the results based on meat-analysis of low risk-of-bias studies. In the consideration of consistency, the lower grade was applied to too strong heterogeneity which disturbed the final interpretation[8]. In the consideration of imprecision, the optimal information size (OIS) for continuous variables would be calculated. If the included studies did not reach OIS or results` 95%CI of the meta-analysis were overlarge, I downgrade the evidence certainty in the concern of impression[8]. The indirectness's grading depended on the eligibility criteria of interpreted studies, especially in the comparison of population[8]. If participants in studies differed from the review`s proposal, the lower grade would be applied. Our downgrading in publication bias depended on the degree of the funnel plot`s asymmetry and statistical test for small-study effect.

**Judgement decision**

*Risk-of bias*

We removed trials with at least some concern of bias in the main interpretation, so we have high certainty in the consideration of risk-of-bias for all outcomes.

*Imprecision*

We set the type-1 error`s level as 0.05 and the power as 80%. Based on one included trial (Asghari 2019[10]) which calculated the appropriate sample size, we choose the post-trial means in the control and zinc intervention arm (183 and 152.1) and their SE for fasting blood glucose to calculate the OIS. The OIS was 45 participants in each arm. None of the outcomes included studies reached this number. But 95% CIs for each outcome were not overly large, we downgraded to moderate certainty in the consideration of imprecision.

*Inconsistency*

There was some concern of inconsistency between T2D and non-specified diabetes patients for serum insulin level by significant subgroup difference. The heterogeneity seemed not to disturb the pooled estimate, since the result of two subgroups and overall were all insignificant. We downgraded our confidence from high to moderate. Other outcomes did not have strong inconsistency and we used the random-effect model. So that, the rest three outcome have high-level certainty in consideration of inconsistency

*Indirectness*

The indirectness consideration for all four outcomes was downgraded to the moderate level because a part of included studies` population did not specified diabetes type and was with complication, which potentially had a discrepancy with the intended T2D population for the review. We did the subgroup analysis and test the group difference for this concern.

*Publication bias*

For the outcome of fasting blood glucose, we downgraded the consideration of publication bias to a moderate level of quality, since its funnel plot was not very symmetrical. The other outcome`s plots maintained fair symmetry so that they were rated high-level certainty.

Overall, the results of fasting blood glucose, HbA1C, HOMA-IR, and serum insulin level had moderate-level confidence in certainty (Tab. S2). Because they had a least one consideration was in moderate certainty evidence.

Table S2: GRADE evaluation of each outcome`s certainty

| Outcomes | Risk of bias | Imprecision | Inconsistency | Indirectness | Publication bias | Overall |
|---|---|---|---|---|---|---|
| Fasting blood glucose | ⊕⊕⊕⊕ High | ⊕⊕⊕ Moderate | ⊕⊕⊕⊕ High | ⊕⊕⊕ Moderate | ⊕⊕⊕ Moderate | ⊕⊕⊕ Moderate |
| HbA1C | ⊕⊕⊕⊕ High | ⊕⊕⊕ Moderate | ⊕⊕⊕⊕ High | ⊕⊕⊕ Moderate | ⊕⊕⊕⊕ High | ⊕⊕⊕ Moderate |
| HOMA-IR | ⊕⊕⊕⊕ High | ⊕⊕⊕ Moderate | ⊕⊕⊕⊕ High | ⊕⊕⊕ Moderate | ⊕⊕⊕⊕ High | ⊕⊕⊕ Moderate |
| Serum insulin level | ⊕⊕⊕⊕ High | ⊕⊕⊕ Moderate | ⊕⊕⊕ Moderate | ⊕⊕⊕ Moderate | ⊕⊕⊕⊕ High | ⊕⊕⊕ Moderate |

**Supplementary material 4: Forest plot of the standardized mean difference of each outcome`s change score between zinc intervention and control arms for low risk-of-bias trials**

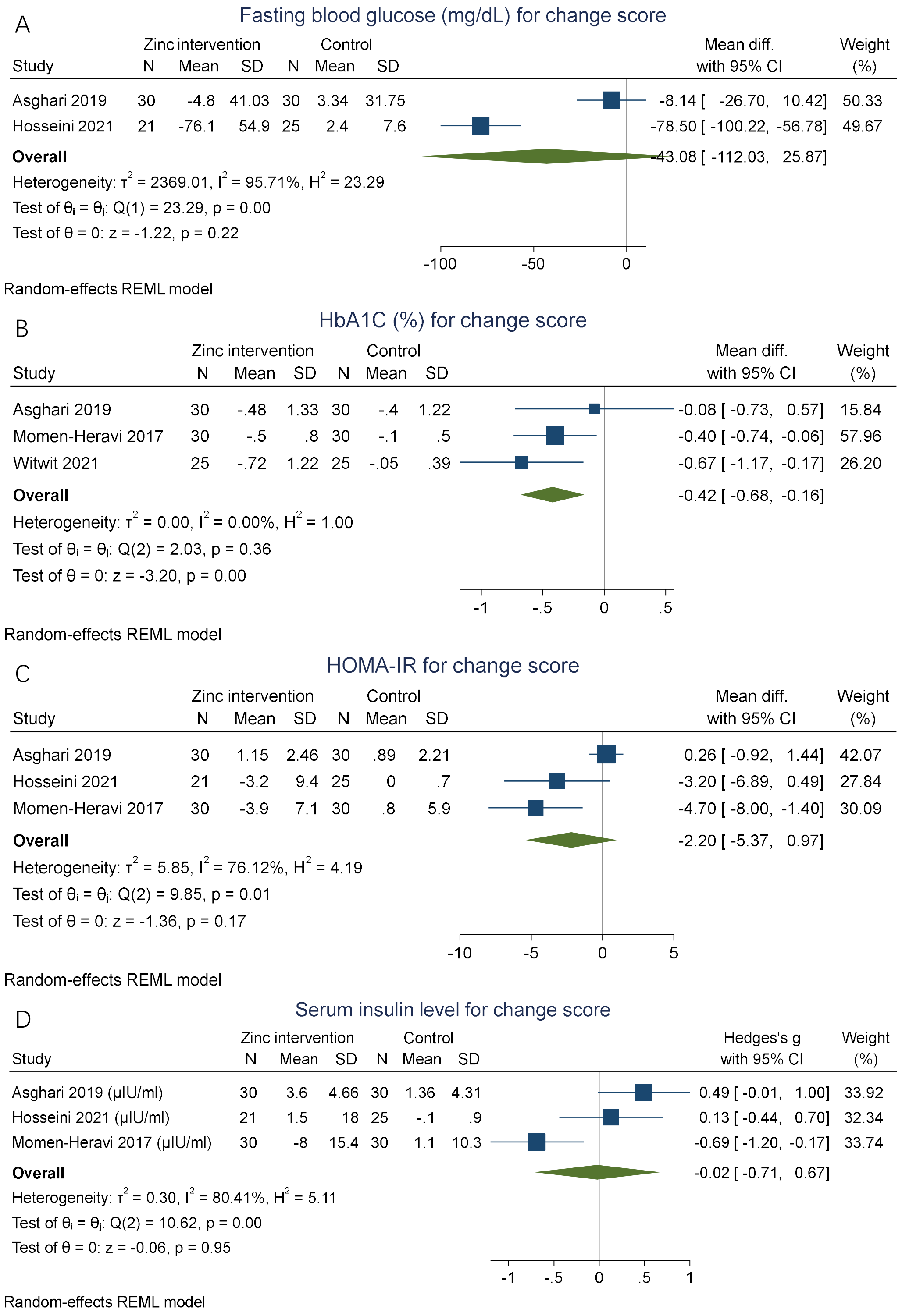

**Supplementary material 5: Post-hoc analysis between zinc intervention and other outcomes**

Some secondary outcomes were found after full-text reading. Without considering the risk-of-bias, three trials reported the post-trial mean of 2h-postprandial blood sugar (Fig. S2). We found zinc intervention`s 2h-postprandial blood sugar was significantly lower than control arm (SMD: -1.11, 95%CI: -1.71, -0.50), with 3 trials at end of the trial. While there was only weak evidence to indicate a difference in the post-trial fasting plasma glucose between zinc intervention and control arms (MD: -13.54, 95%CI: -48.5, 21.41), with 2 trials. These meta-analyses used the random-effect model.

One study reported a significant decrease in random blood glucose between pre-trial and post-trial in the zinc intervention arm and this decrease was stronger than the change in the control arm[11]. Another study indicated that serum homocysteine levels also reduced significantly in the zinc intervention arm. Homocysteine is a homologue of the amino acid cysteine, which was found to correlate with insulin resistance through hyperhomocysteinemia and other metabolic mechanism[12,13].

Figure S2: Forest plot of the standardized mean difference of post-trial 2h-postprandial blood sugar and fasting plasma glucose between zinc intervention and control arms

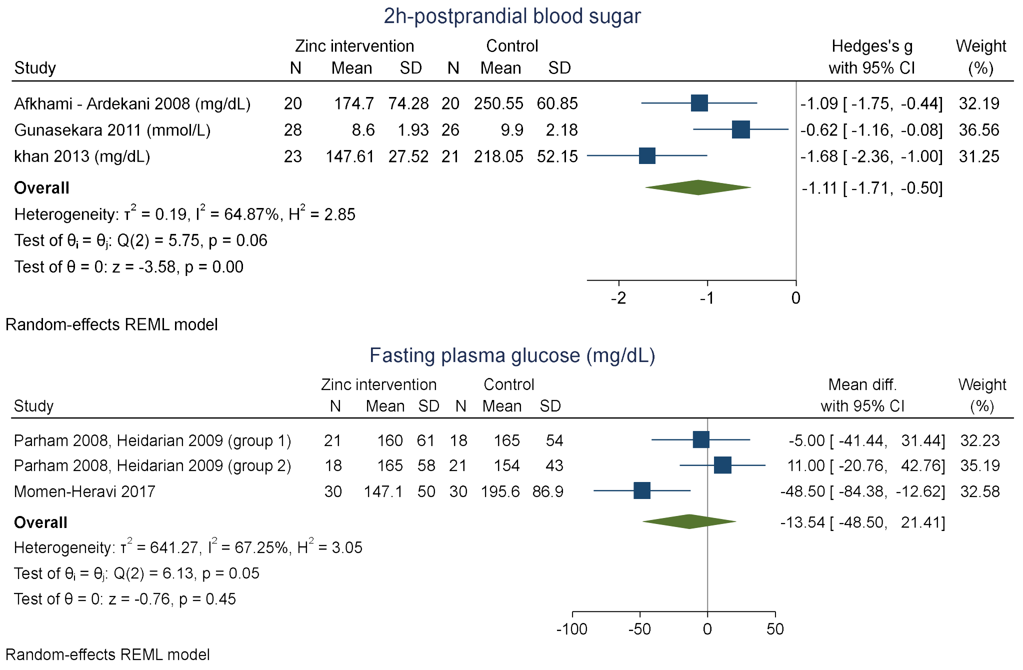

**Supplementary material 6: Forest plot of the mean difference or standardized mean difference of post-trial outcomes between zinc intervention and control arm for all trials.**

Fasting blood glucose (mg/dL)
in all trials

| Study | Zinc intervention N | Zinc intervention Mean | Zinc intervention SD | Control N | Control Mean | Control SD | Mean diff. with 95% CI | Weight (%) |
|---|---|---|---|---|---|---|---|---|
| Afkhami - Ardekani 2008 | 20 | 134.25 | 57.81 | 20 | 159.5 | 27.35 | -25.25 [ -53.28, 2.78] | 6.65 |
| Asghari 2019 | 30 | 152.1 | 45.2 | 30 | 183 | 57.6 | -30.90 [ -57.10, -4.70] | 7.25 |
| Gunasekara 2011 | 28 | 106.02 | 18.18 | 26 | 117.18 | 20.34 | -11.16 [ -21.43, -0.89] | 15.62 |
| Hosseini 2021 | 21 | 132 | 47.9 | 25 | 184.1 | 52.2 | -52.10 [ -81.28, -22.92] | 6.30 |
| khan 2013 | 23 | 113.74 | 23.38 | 21 | 154.95 | 23.38 | -41.21 [ -55.04, -27.38] | 13.35 |
| Matter 2020 | 39 | 116.9 | 4.6 | 38 | 144.5 | 22.9 | -27.60 [ -34.93, -20.27] | 17.41 |
| Naghizadeh 2018 | 23 | 162.95 | 28.22 | 22 | 200.27 | 55.9 | -37.32 [ -63.02, -11.62] | 7.43 |
| Nazem 2019 | 35 | 121.94 | 21.15 | 35 | 149.43 | 25.84 | -27.49 [ -38.55, -16.43] | 15.11 |
| Pérez 2018 | 13 | 124 | 21.1 | 15 | 130.4 | 26.7 | -6.40 [ -24.43, 11.63] | 10.88 |
| **Overall** | | | | | | | -26.87 [ -35.74, -18.00] | |

Heterogeneity: $\tau^2 = 103.66$, $I^2 = 66.01\%$, $H^2 = 2.94$

Test of $\theta_i = \theta_j$: Q(8) = 21.36, p = 0.01

Test of $\theta = 0$: z = -5.94, p = 0.00

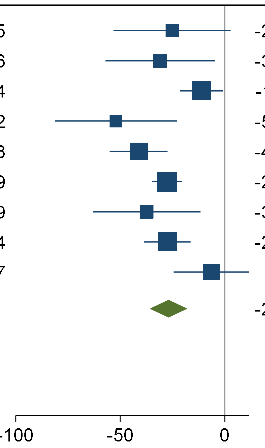


Random-effects REML model

HbA1C (%)
in all trials

| Study | Zinc intervention N | Zinc intervention Mean | Zinc intervention SD | Control N | Control Mean | Control SD | Mean diff. with 95% CI | Weight (%) |
|---|---|---|---|---|---|---|---|---|
| Afkhami - Ardekani 2008 | 20 | 7.35 | 1.62 | 20 | 7.46 | .73 | -0.11 [ -0.89, 0.67] | 10.22 |
| Asghari 2019 | 30 | 8.4 | 1.3 | 30 | 8.6 | 1.6 | -0.20 [ -0.94, 0.54] | 10.62 |
| Gunasekara 2011 | 28 | 6.84 | .91 | 26 | 8.2 | 1.28 | -1.36 [ -1.95, -0.77] | 12.11 |
| khan 2013 | 23 | 6.91 | .67 | 21 | 8.61 | .92 | -1.70 [ -2.17, -1.23] | 13.27 |
| Momen-Heravi 2017 | 30 | 7.3 | 1.1 | 30 | 7.8 | .6 | -0.50 [ -0.95, -0.05] | 13.51 |
| Naghizadeh 2018 | 23 | 8.22 | 1.57 | 22 | 9.02 | 1.27 | -0.80 [ -1.64, 0.04] | 9.68 |
| Nazem 2019 | 35 | 7.35 | .57 | 35 | 7.56 | .55 | -0.21 [ -0.47, 0.05] | 15.08 |
| Parham 2008, Heidarian 2009 (group 1) | 21 | 7.2 | 1.4 | 18 | 7.4 | 1.8 | -0.20 [ -1.20, 0.80] | 8.23 |
| Parham 2008, Heidarian 2009 (group 2) | 18 | 7.2 | 1.8 | 21 | 7.5 | 1.8 | -0.30 [ -1.43, 0.83] | 7.27 |
| **Overall** | | | | | | | -0.64 [ -1.05, -0.22] | |

Heterogeneity: $\tau^2 = 0.28$, $I^2 = 77.12\%$, $H^2 = 4.37$

Test of $\theta_i = \theta_j$: Q(8) = 39.53, p = 0.00

Test of $\theta = 0$: z = -3.03, p = 0.00

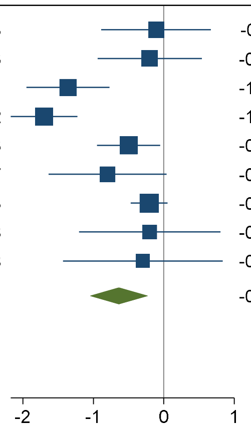


Random-effects REML model

**Supplementary material 7: Association between calcium supplementary and diabetes outcome in two-sample MR**

| Outcome | N. of SNP | IVW coefficient | P-value |
|---|---|---|---|
| Fasting glucose | 3 | -0.32 | 0.62 |
| HbA1C | 3 | 0.36 | 0.56 |
| HOMA-IR | 3 | -0.65 | 0.37 |
| Insulin level | 3 | 0.09 | 0.83 |

**Supplementary material 8: Association between zinc supplementary and hair color in two-sample MR**

| hair colour | N. of SNP | IVW coefficient | P-value |
|---|---|---|---|
| Blonde | 3 | -0.20 | 0.27 |
| Red | 3 | -0.14 | 0.46 |
| Light brown | 3 | 0.16 | 0.58 |
| Dark brown | 3 | 0.03 | 0.94 |
| Black | 3 | 0.16 | 0.18 |
| other | 3 | 0.00 | 0.95 |
